# Gestational Hypertension in Adolescent Mothers: A 2016-2022 Trend Analysis

**DOI:** 10.1101/2024.08.20.24312329

**Authors:** Oluwapelumi Oloyede, Lulu Xu, Omolola Adepoju

## Abstract

Hypertensive disorders in pregnancy (HDPs) significantly contribute to maternal and fetal complications, particularly in adolescent pregnancies. This study examines the prevalence and predictors of gestational hypertension (gHTN) among U.S. adolescents between 2016 and 2022, using data from the CDC’s Birth Data Files. The analysis included various maternal factors, such as age, race, education, BMI, prenatal care, and participation in the Women, Infants, and Children (WIC) Nutritional Program. Logistic Regression and Random Forest models were employed to evaluate these predictors, with Random Forest showing superior predictive performance. The study found that gHTN prevalence increased from 6.72% in 2016 to 9.51% in 2022, with BMI, month prenatal visits began, WIC participation, and race emerging as key predictors. The findings highlight the importance of early prenatal care and targeted support for adolescents to manage gHTN, emphasizing the need for interventions that address modifiable risk factors such as BMI and access to nutritional programs. This research underscores the critical need for continued efforts to mitigate the rising trend of gHTN in adolescent pregnancies and improve maternal and fetal outcomes in this vulnerable population. Future studies should focus on identifying additional predictors and tailoring interventions to meet the unique needs of adolescent mothers.

## Introduction

Hypertensive disorders in pregnancy (HDPs) are among the leading causes of maternal and fetal complications throughout pregnancy and delivery. These complications determine the maternal course of when a mother may deliver and the means by which she will do so. HDPs are known to cause complications such as myocardial infarctions, stroke, preterm labor, and placental abruption. About 8% of singleton pregnancies develop hypertensive disorders in pregnancy, and from 1989 to 2020, the prevalence of HDPs increased from 2.79% to 8.22% [1]. Collectively, HDPs often present with a wide range of variability, including chronic hypertension, gestational hypertension, preeclampsia, and chronic hypertension with superimposed preeclampsia.

Gestational hypertension (gHTN) is one of the most commonly reported HDPs[2]. The American College of Obstetricians and Gynecologists (ACOG) defines gHTN as blood pressure readings greater than or equal to 140mmHg systolic or 90mmHg diastolic on two separate occasions at least four hours apart after 20 weeks of pregnancy[3]. Costs associated with gHTN pregnancies are markedly higher than in pregnancies without hypertension ($12,784 vs. $8,854)[4]. These trends are more concerning in adolescent mothers, with earlier work suggesting that adolescent mothers face higher risks of hypertensive diseases of pregnancy, such as eclampsia and infections, compared to women aged 20–24 years[5]. While the Centers for Disease Control and Prevention (CDC) reports that overall teen births are declining, the US teen birth rate remains higher than in many other affluent countries[6]. The rate of adolescent pregnancies warrants a discussion about how this population may be affected by alarming diagnoses that can lead to adverse outcomes. Despite significant efforts addressing fetal and maternal outcomes, particularly focusing on preeclampsia and eclampsia, our literature review identified a paucity of studies on other HDPs, such as gHTN, that affect adolescent pregnancies.

Despite the widespread recognition of the issue of HDPs, there remains a gap in our understanding of HDPs in adolescent mothers. Most adolescent mothers are often first-time mothers or nulliparous, and specifically, the literature reports that nulliparous women may require low thresholds to identify their hypertensive states[7]. Understanding factors contributing to the increasing prevalence of gHTN in adolescent pregnancies can help in developing strategies to manage this population and prevent further adverse life events. This retrospective study evaluates patterns of gHTN between 2016-2022, among US adolescents.

## METHODS

### Data

This study used data from Birth Data Files obtained from the Centers for Disease Control and Prevention (CDC) Vital Statistics division, from 2016-2022. These data files are part of the National Vital Statistics System (NVSS) and are collected through a cooperative effort between the National Center for Health Statistics (NCHS) and all U.S. jurisdictions, including the 50 states, the District of Columbia, and U.S. territories.[8] This dataset has been used in previous studies to explore maternal and fetal outcomes. In this study, we examined gHTN in adolescents. The analysis focused on a subset of records with birth moms less than 20 years of age.

### Measurement

The dependent variable was a binary flag indicating whether a patient had gestational diabetes (yes/no). Predictor variables included Mother’s Age, Mother’s Race, Mother’s Ethnicity, Marital Status, Mother’s Education (“1”: 8th grade or less, “2”: High school graduate, “3”: Some college, “4” Bachelor’s degree), Live Birth Order (“1-7”: Number of live birth order, “8”: 8 or more live births), Month Prenatal Care Began (“1”: 1st to 3rd month, “2”: 4th to 6th month, “3”: 7th to final month, “4” No prenatal care), Number of Prenatal Visits (“00-98”: Number of prenatal visits), Receipt of the Woman, Infant and Children (WIC) (“Y”: Yes, “N”: No), Body Mass Index (BMI) (“1”: Underweight <18.5, “2”: Normal 18.5-24.9, “3”: Overweight 25.0-29.9, “4”: Obesity I 35.0-34.9, “5”: Obesity II 35.0-39.9, “6” Extreme Obesity III ≥ 40.0), and a binary flag indicating whether the mother had Pre-pregnancy Hypertension. These variables were included based on empirical literature evidence on their importance on pregnancy outcomes.

### Data Loading and Initial Processing

The dataset included 1.048 million observations and 13 features, comprising 11 predictors, 1 response variable, and a “YEAR” variable. Initial checks confirmed the categorical nature of all features. Exploratory Data Analysis (EDA) was conducted to understand the distribution of the variables, including evaluating the distribution of the response variable, gHTN. The distribution analysis revealed a significant class imbalance in the response variable, with 91.87% ‘No’ and 8.13% ‘Yes’. Correlation analysis using Cramer’s V indicated no strong correlations between the predictors. These findings informed our data processing and modeling approach.

### Data Preprocessing

We separated the dataset by year to examine changes in predictors associated with gHTN between 2016 and 2022. From previous distribution analysis, we found that the response variable, gHTN, exhibited a significant class imbalance.[9] To address this issue, the Synthetic Minority Over-sampling Technique (SMOTE) was applied to generate synthetic samples and balance the classes.[10] This step was crucial for preventing model bias, improving accuracy and recall, and enabling more effective learning. The balanced dataset was then split into training (80% of the data) and test sets (20% of the data) using Python’s **train_test_split** function from **sklearn**.**model_selection** [11] to facilitate model building and evaluation

### Model Building

For both interpretability and robustness, and to provide a comprehensive analysis of predictors’ impacts on gHTN while ensuring reliable predictions, we chose two predictive models: Logistic Regression[12] and Random Forest.[13] These models were used to build and compare predictions for the two selected years, 2016 and 2022. Although the years 2017 to 2021 were analyzed, they were not included in the direct model comparison because a longer time span was needed to observe clear changes, making the selected years more suitable for comparison. These middle years were primarily used to ensure that the trends observed in 2016 and 2022 were not anomalies but reflected consistent patterns across the period. By focusing on 2016 and 2022, we aimed to contrast periods over a long span of years to observe how variables impacted changes in gHTN Logistic regression was chosen for its simplicity and interpretability. This model estimates the probability of a binary outcome based on one or more predictor variables. Its coefficients provide insights into the relationship between each predictor and the response variable, making it a good baseline model for understanding predictor impacts. Random Forest was selected for its robustness and ability to handle complex interactions between variables, particularly given our study’s categorical data. This ensembles learning method to construct multiple decision trees during training and outputs the mode of the classes (classification) of the individual trees. Random forests can model non-linear relationships and are less prone to overfitting compared to a single decision tree.

Model performance was evaluated based on accuracy, precision, recall, and F1-score to ensure a comprehensive assessment of each model’s capabilities in predicting gHTN[14]. The impact of predictors was assessed using logistic regression coefficients and random forest feature importance scores, providing a detailed understanding of how each predictor influences the risk of gHTN.

## Results

Table 1 shows the descriptive characteristics of the study sample. The prevalence of gHTN ranged from 6.72% in 2016 to 9.51% in 2022. The prevalence of gHTN ranged from 6.72% in 2016 to 9.51% in 2022. Respondents who identified as White experienced a slight decrease in gHTN prevalence, from 69.88% in 2016 to 67.43% in 2022, while gHTN prevalence nearly decreased by half for Asian respondents, from 1.45% in 2016 to 0.87% in 2022. However, gHTN prevalence slightly increased for both Black and AIAN respondents during this period, from 22.35% in 2016 to 24.46% in 2022 and 1.84% in 2016 to 1.96% in 2022 respectively. Hispanic respondents experienced a small decrease in gHTN prevalence from 35.93% in 2016 to 35.74% in 2022, along with Non-Hispanic White and AIAN respondents, from 37.76% in 2016 to 35.96% in 2022 and 1.51% in 2016 to 1.42% in 2022 respectively. While Non-Hispanic Asian respondents experienced the sharpest decrease, from 0.90% in 2016 to 0.60% in 2022, gHTN prevalence only increased for Non-Hispanic Black respondents, from 19.90% in 2016 to 21.61% in 2022. Regarding marital status, gHTN prevalence decreased by a-fifth for married respondents, from 10.88% in 2016 to 8.10% in 2022, and increased for respondents who were not married, from 89.12% in 2016 to 91.90% in 2022. When probed for education status, gHTN prevalence increased for both respondents who reported an education of 8^th^ Grade or less, from 4.22% in 2016 to 5.82% in 2022, and 9^th^-12^th^ Grade without a diploma, from 85.28% in 2016 to 86.23% in 2022. However, gHTN prevalence decreased for respondents who reported having a High School Graduate diploma/GED, from 10.48% in 2016 to 7.95% in 2022. No change in prevalence was reported for respondents reporting some college education (no degree), with available data reporting 0.02% from 2016 to 2017.

**Table 1:**
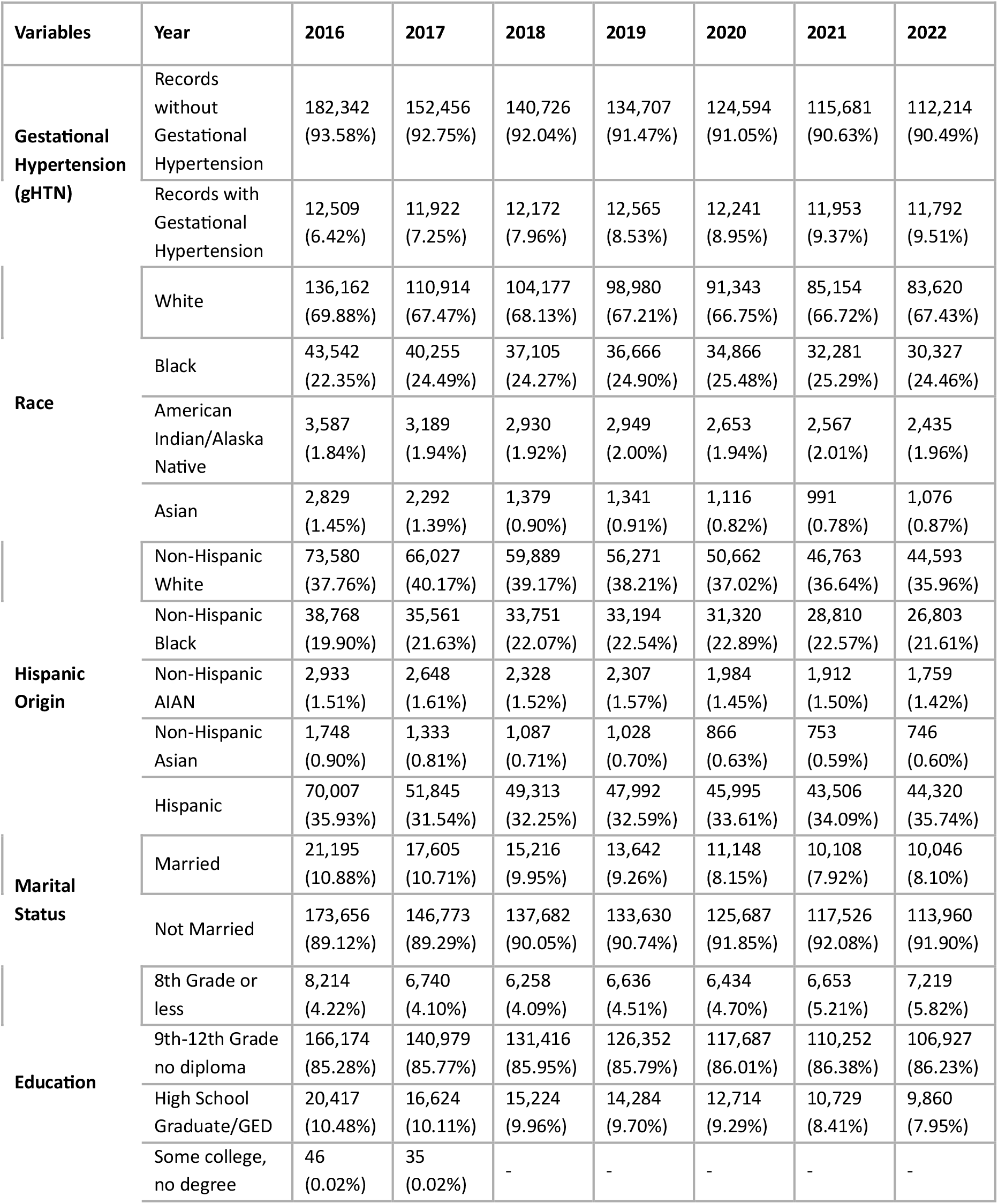
Descriptive Characteristics of Study Sample by Year, n %.

### Comparative Performance Analysis

The comparative performance analysis, described in Table 2, demonstrates slight improvements in model performance over time. Both models showed modest enhancements in accuracy from 2016 to 2022. For instance, the accuracy of the Logistic Regression model increased marginally from 0.63 in 2016 to 0.64 in 2022, while the Random Forest model’s accuracy rose slightly from 0.71 in 2016 to 0.72 in 2022. These small increases indicate a gradual improvement in model reliability and predictive capabilities over the years.

**Table 2:** Comparative Performance of Logistic Regression and Random Forest Models for Predicting Outcomes in 2016 and 2022.

| Year | Model | Accuracy | Precision Class 0 | Precision Class 1 | Recall Class 0 | Recall Class 1 | F1-Score Class 0 | F1-Score Class 1 | Support Class 0 | Support Class 1 |
| --- | --- | --- | --- | --- | --- | --- | --- | --- | --- | --- |
| 2016 | Logistic Regression | 0.63 | 0.60 | 0.69 | 0.80 | 0.46 | 0.68 | 0.55 | 36,497 | 36,440 |
| 2016 | Random Forest | 0.71 | 0.71 | 0.72 | 0.72 | 0.70 | 0.72 | 0.71 | 36,497 | 36,440 |
| 2022 | Logistic Regression | 0.64 | 0.60 | 0.70 | 0.79 | 0.48 | 0.68 | 0.57 | 22,323 | 22,563 |
| 2022 | Random Forest | 0.72 | 0.71 | 0.73 | 0.73 | 0.70 | 0.72 | 0.71 | 22,323 | 22,563 |

Additionally, the Random Forest model consistently outperformed the Logistic Regression model across all metrics for both years. In 2022, the Random Forest model achieved a precision for Class 1 of 0.73, compared to the Logistic Regression model’s 0.70. The F1-score for Class 1 was 0.71 for the Random Forest model, significantly higher than the 0.57 achieved by the Logistic Regression model. This consistent superior performance highlights the robustness and effectiveness of the Random Forest model in predicting outcomes.

These results emphasize the importance of model selection in predictive analytics and demonstrate the Random Forest model’s superior capability in handling complex datasets and providing reliable predictions.

### Analysis of Feature Impact

Figure 1 presents a comparative analysis of the impact of various features on the prediction of gHTN using Logistic Regression and Random Forest models for the years 2016 and 2022. Most features exhibit slight variations in their coefficients and importance scores. The Logistic Regression model generally assigns higher numerical values to coefficients, while the Random Forest model distributes importance more evenly across features. This distribution suggests that the Random Forest model captures a broader array of interactions between features.

**Figure 1.**
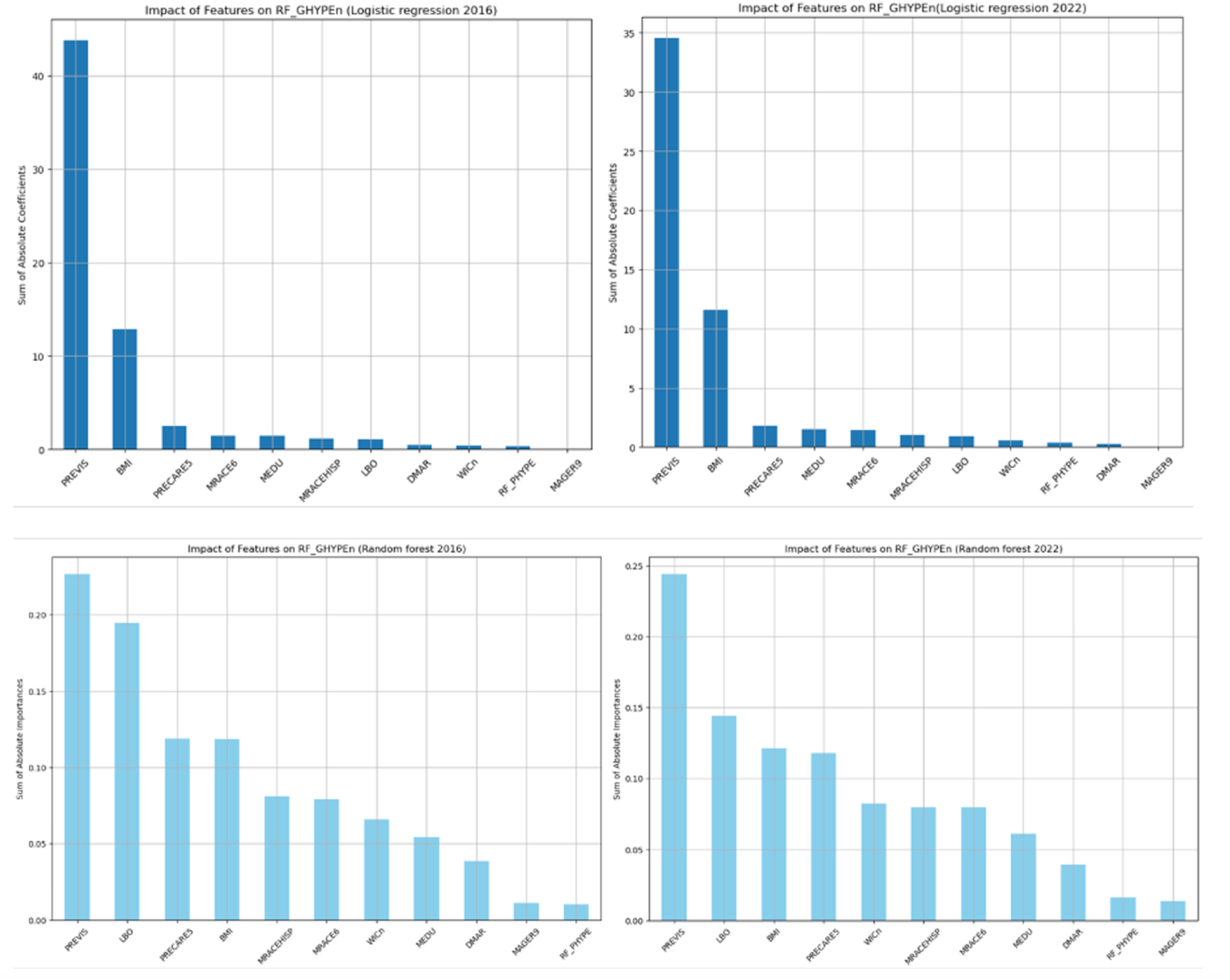
Comparative Analysis of Feature Impact of Gestational Hypertension Models: Logistic Regression vs. Random Forest (2016 & 2022)

The analysis reveals that the number of prenatal visits is the most significant feature across both models and years, indicating its strong and stable correlation with a diagnosis of gHTN. In addition, BMI also consistently shows high coefficients and importance scores. In the Logistic Regression model, BMI holds the second-highest place in importance, underscoring its crucial role in predicting gHTN. Similarly, in the Random Forest model, BMI maintains a high importance score, ranking third in 2016 and fourth in 2022, indicating its significant impact on the outcome.

In the Logistic Regression model, the rest of the factors all had minimal importance compared to number of prenatal visits and BMI. In contrast, the Random Forest model shows that several other factors have relatively significant importance over both years. For example, the order of live births rose to importance as the second most important factor, even though its absolute impact on predicting gHTN has diminished over time. The importance of month prenatal care began and BMI factors remained relatively high and stable from 2016 to 2022. Additionally, the importance of receipt of WIC increased over the years, indicating that participation in nutritional programs has become increasingly important in predicting gHTN outcomes.

These results highlight the changing dynamics of feature importance over time and underscore the need for continuous updates to the models to ensure they remain effective in predicting gHTN.

## Discussion

This study examined trends contributing to gHTN in adolescents, and how those trends changed between 2016 and 2022. Based on the most important features in both models, we found BMI, month prenatal care started and race/ethnicity to be the strongest features associated with gHTN in adolescents in 2016, compared to BMI, month prenatal care began, receipt of WIC, and Education in 2022. The count of prenatal visits remained strongly associated with gHTN over time. While we only observed modest changes in these trends over time, studies of this nature can be valuable for future healthcare professionals and policies aimed at the management of gHTN in this understudied adolescent subpopulation.

From the beginning of pregnancy, the significance of prenatal visits becomes paramount for monitoring potential risks and complications. Given the well-established association between adolescent pregnancies and adverse fetal and maternal outcomes, prenatal visits, as our models showed, emerged as the most significant feature across various years and models. The positive association between prenatal visits and the management of gHTN in adolescents suggests the healthcare system’s role in providing additional access points for adolescents with a diagnosis of gHTN. Highlighting such successes is crucial to this field, as it validates existing obstetrics health strategies but also balances the problem-focused narratives that exist in research today. The association also emphasizes a bigger point of adherence to medical appointments and following healthcare recommendations.

Obesity in pregnancy is well-documented to have a strong association with pregnancy complications, and our finding on BMI being one of the strongest predictors aligns with these earlier studies. A study involving almost 4000 pregnant women investigated the impact of pre-pregnancy BMI and gestational weight gain on adverse pregnancy outcomes, revealing that pre-pregnancy BMI was associated with an increased risk of preeclampsia, gHTN, gestational diabetes, and macrosomia.[15] Furthermore, a prospective cross-sectional study involving 365 singleton adolescent pregnancies aged between 16 and 20 years found that overweight/obese adolescents exhibited a higher risk of cesarean delivery, preeclampsia, and small for gestational age.[16] These findings emphasize the importance of addressing pre-pregnancy BMI as a potential modifiable risk factor in adolescent pregnancies. The potential maternal and fetal outcomes among overweight/obese adolescents highlight the need for targeted interventions and preconception counseling to optimize outcomes in this vulnerable population.

The timing of the first prenatal visit was also significantly associated with gestational hypertension and remained consistently significant from 2016 to 2022. Previous research has emphasized the importance of early prenatal care and its positive impact on maternal outcomes. Late initiation of prenatal care is widely recognized as a risk factor for HDPs, including preeclampsia and gHTN.[17] Early prenatal visits, particularly those occurring in the first trimester, can act as a protective factor by preventing disease progression and improving maternal and fetal outcomes. This finding underscores the critical need to address the barriers that prevent adolescent mothers from accessing timely prenatal care. A large-scale survey involving 31,642 women revealed that many women expressed a desire for earlier prenatal visits but faced significant obstacles. These barriers included the unavailability of appointments, insurance issues, financial constraints, and delayed recognition of pregnancy.[18] Addressing these financial and structural barriers is essential for supporting adolescent mothers in receiving the care they need. Furthermore, education plays a crucial role in influencing the timing of pregnancy recognition and the initiation of prenatal care. Comprehensive education efforts, particularly targeted at adolescents, are necessary to ensure early engagement with healthcare services, ultimately reducing the risk of adverse outcomes associated with gestational hypertension.

The increased use of the Women, Infants, and Children (WIC) Nutritional Program shows its growing importance in predicting and potentially reducing gestational hypertension. This finding highlights the need to screen expectant mothers for WIC assistance and address their specific needs. It is especially important to encourage adolescents to use these resources. Additionally, this research emphasizes the need for health equity within WIC, ensuring that all pregnant women have access to nutritional support, regardless of their socioeconomic status. WIC has been shown to improve maternal and fetal outcomes, leading to lower rates of preterm birth and infant mortality among mothers on Medicaid who receive WIC benefits[19]. This calls for better outreach and support for the WIC program, so more expectant mothers can benefit from it. Future research should look at which parts of WIC are most helpful and explore other factors that affect its effectiveness.

It is widely acknowledged that race has historically played a significant role in women’s health. The impact of race was therefore anticipated in our results, yet it remains a critical focus in our research, highlighting how current practices disproportionately affect women of different racial and ethnic backgrounds. Racial disparities can stem from various factors, including limited access to quality healthcare, systemic bias within medical institutions, socioeconomic inequalities, and differences in social determinants of health. Patients will benefit significantly when providers are informed by research like this, enabling them to offer more equitable care to all individuals.

There are many limitations to be considered when interpreting these findings. This analysis examines associations and does not insinuate causation. Confounding variables, such as the impact of COVID-19, may have influenced outcomes and were not fully accounted for in the analysis. While random forest models offer advantages, such as handling non-linear relationships, they may not always outperform logistic regression in all scenarios, suggesting potential room for methodological refinement in future studies. Despite these limitations, there are many strengths, including the large sample size, which provides statistical power and enhances the generalizability of findings. Additionally, the study spanned a significant time period and utilized a national dataset, enhancing the scope of the analysis.

## CONCLUSION

In conclusion, this study explores the multifaceted predictors and trends of gestational hypertension (gHTN) in adolescent pregnancies, extending beyond the commonly discussed racial disparities. We noted a steady rise in gHTN in this high-risk group, emphasizing the need for targeted monitoring and intervention strategies. The challenges of psychological and physical health can be mitigated through better prenatal care, social support, and education to prevent adverse outcomes associated with gHTN.

The role of prenatal visits is crucial in managing gHTN and sheds light on the importance of proactive care. Other factors such as BMI and participation in the WIC Nutritional Program highlight the importance of comprehensive support programs for expecting adolescents. These are modifiable risk factors that can be used as a basis for health strategies to improve maternal and fetal outcomes in this vulnerable population.

Future research is needed to explore additional predictors of gHTN to tailor interventions and address the unique needs of this high-risk group.

## Data Availability

The data that support the findings of this study are available in the CDC's Vital Statistics Online Data Repository at https://www.cdc.gov/nchs/data_access/VitalStatsOnline.htm#Births. These data were derived from the following public domain resources: the CDC's Birth Data Files, accessible through the National Center for Health Statistics (NCHS).

https://www.cdc.gov/nchs/data_access/vitalstatsonline.htm

